# Anti-membrane and anti-spike antibodies are long-lasting and together discriminate between past COVID-19 infection and vaccination

**DOI:** 10.1101/2021.11.02.21265750

**Authors:** Maya F. Amjadi, Ryan R. Adyniec, Srishti Gupta, S. Janna Bashar, Aisha M. Mergaert, Katarina M. Braun, Gage K. Moreno, David H. O’Connor, Thomas C. Friedrich, Nasia Safdar, Sara S. McCoy, Miriam A. Shelef

**Author notes:** **Corresponding author:** Miriam A. Shelef, 4130 UW Medical Foundation Centennial Building, 1685 Highland Avenue, Madison, WI 53705.

## Abstract

The consequences of past COVID-19 infection for personal health and long-term population immunity are only starting to be revealed. Unfortunately, detecting past infection is currently a challenge, limiting clinical and research endeavors. Widely available anti-SARS-CoV-2 antibody tests cannot differentiate between past infection and vaccination given vaccine-induced anti-spike antibodies and the rapid loss of infection-induced anti-nucleocapsid antibodies. Anti-membrane antibodies develop after COVID-19, but their long-term persistence is unknown. Here, we demonstrate that anti-membrane IgG is a sensitive and specific marker of past COVID-19 infection and persists at least one year. We also confirm that anti-receptor binding domain (RBD) Ig is a long-lasting, sensitive, and specific marker of past infection and vaccination, while anti-nucleocapsid IgG lacks specificity and quickly declines after COVID-19. Thus, a combination of anti-membrane and anti-RBD antibodies can accurately differentiate between distant COVID-19 infection, vaccination, and naïve states to advance public health, individual healthcare, and research goals.

## Introduction

The cardinal features and challenges of the COVID-19 pandemic are changing. Initially, the pandemic was defined by acute SARS-CoV-2 infections in an immunologically naive population. Now, although acute infections continue to devastate many communities, population immunity from vaccination and past infection are rising. However, the differences in protection due to the two types of immunity remain only partially defined. Further, medium and long term health consequences of COVID-19 are being revealed ^1^, but these can be difficult to diagnose and study particularly for individuals without viral test results during acute illness. To tackle these new challenges, scientists and clinicians need easily deployable testing strategies to differentiate between past infection and vaccination.

Serologic antibody detection is a standard method for grossly evaluating humoral immunity in response to vaccination and infection for research and clinical purposes. For COVID-19, the vast majority of serologic tests detect antibodies against the SARS-CoV-2 spike protein or nucleocapsid protein ^2^. However, anti-nucleocapsid antibodies can decline to the point of seronegativity just months after infection, making them an unreliable marker of distant infection ^3-5^. Anti-spike antibodies persist at least a year post-infection ^6,7^, but serologic testing for anti-spike or receptor binding domain (RBD) of spike antibodies cannot differentiate between past COVID-19 infection and vaccination with any of the vaccines approved in the United States, all of which rely upon the spike protein ^8-10^. The membrane protein is the most abundant SARS-CoV-2 structural protein. However, serological testing of anti-membrane protein antibodies is in its infancy. Antibodies against membrane antigens are present days to months after SARS-CoV-2 infection ^11-13^, but it is unknown how long they persist. Thus, there are currently no widely available serologic tests that uniquely and accurately detect past COVID-19 infection, limiting research and public health efforts.

## Results and Discussion

To identify SARS-CoV-2 antigens that could be used in serologic tests to reliably discriminate between past infection, past vaccination, and naïve states, we quantified antibody levels against the RBD of spike using a commercially available chemiluminescent assay and against the nucleocapsid and membrane peptides that we previously identified as best able to discriminate between COVID-19 convalescent and naive subjects by ELISA ^11^. We evaluated the sensitivity and specificity of these tests using sera from naïve subjects (never vaccinated against or infected by SARS-CoV-2 collected prior to 2019) and COVID-19 convalescent subjects 5 weeks post-symptom resolution. We then tested sera from the following subjects: naïve, vaccinated with no known COVID-19, COVID-19 convalescent with sera collected from at least 3 of 4 timepoints post-symptom resolution (5 weeks, 3 months, 6 months, 12 months, all initially unvaccinated and most vaccinated at 12 months), and vaccinated against SARS-CoV-2 with sera collected during subsequent breakthrough COVID-19. Basic clinical and demographic information is provided in Table 1.

**Table 1.** Basic Clinical and Demographic Data for Subjects.

| Cohort | n | Age | Sex (%) |  |  | Days since 1<sup>st</sup> vaccine |
| --- | --- | --- | --- | --- | --- | --- |
|  |  | Median (IQR) | F | M | NB | Median (IQR) |
| Naïve | 60 | 49 (37, 63) | 65 | 35 | 0 | - |
| Vaccinated | 21 | 41 (33, 56) | 90 | 5 | 5 | 186 (170, 206) |
| COVID-19 Convalescent |  |  |  |  |  |  |
| 5 weeks | 104 | 48 (33, 58) | 58 | 42 | 0 | - |
| 3 months | 101 | 45 (33, 58) | 56 | 44 | 0 | - |
| 6 months | 97 | 47 (34, 59) | 60 | 40 | 0 | - |
| 12 months, unvaccinated | 21 | 44 (38, 58) | 48 | 52 | 0 | - |
| 12 months, vaccinated | 77 | 50 (34, 58) | 60 | 40 | 0 | 73 (40, 98) |
| Vaccinated with breakthrough COVID-19 | 20 | 35 (29, 41) | 80 | 15 | 5 | 125 (104, 154) |
| Post-breakthrough, 8 weeks | 3 | 47 (35, 57) | 67 | 33 | 0 | 173 (153, 185) |

Focusing first on anti-RBD Ig, we calculated an AUC of 0.973 (Wald 95% CI 0.945, 0.995) for the chemiluminescent assay with a sensitivity of 90% and specificity of 97% (Figure 1A). We then found that COVID-19 convalescent and vaccinated individuals had significantly higher anti-RBD Ig than naïve subjects, with no significant difference in antibody levels or percent seropositivity between vaccinated subjects and unvaccinated convalescent subjects (Figure 1B and 1C). In a paired analysis of the 88 subjects who provided serum at all four timepoints, anti-RBD Ig levels were statistically different between timepoints (p<0.0001), but this difference seems unlikely to be biologically meaningful given the medians at 5 weeks (5.35), 3 months (4.63), and 6 months (4.95) post-COVID-19 symptom resolution. Further, the percent of seropositive subjects at 5 weeks (91%) versus 6 months (88%) was not significantly different. Because only 21/100 convalescent subjects remained unvaccinated at 12 months, this timepoint was not compared to the 5 week timepoint in either analysis. Not surprisingly, anti-RBD Ig levels in 12 month convalescent subjects were significantly higher for those who received at least one dose of a vaccine compared with no vaccine (Figure 1B and Supplementary Figure 1B). Finally, all vaccinated subjects, with or without past or breakthrough infections, were seropositive for anti-RBD Ig, with no significant differences in antibody levels between vaccinated subjects with or without breakthrough infections. Overall, these data suggest that anti-RBD antibodies are detectable in the vast majority of vaccinated and COVID-19 convalescent subjects even a year after infection, but cannot differentiate between past infection, vaccination, and vaccination with breakthrough infection.

**Figure 1.**
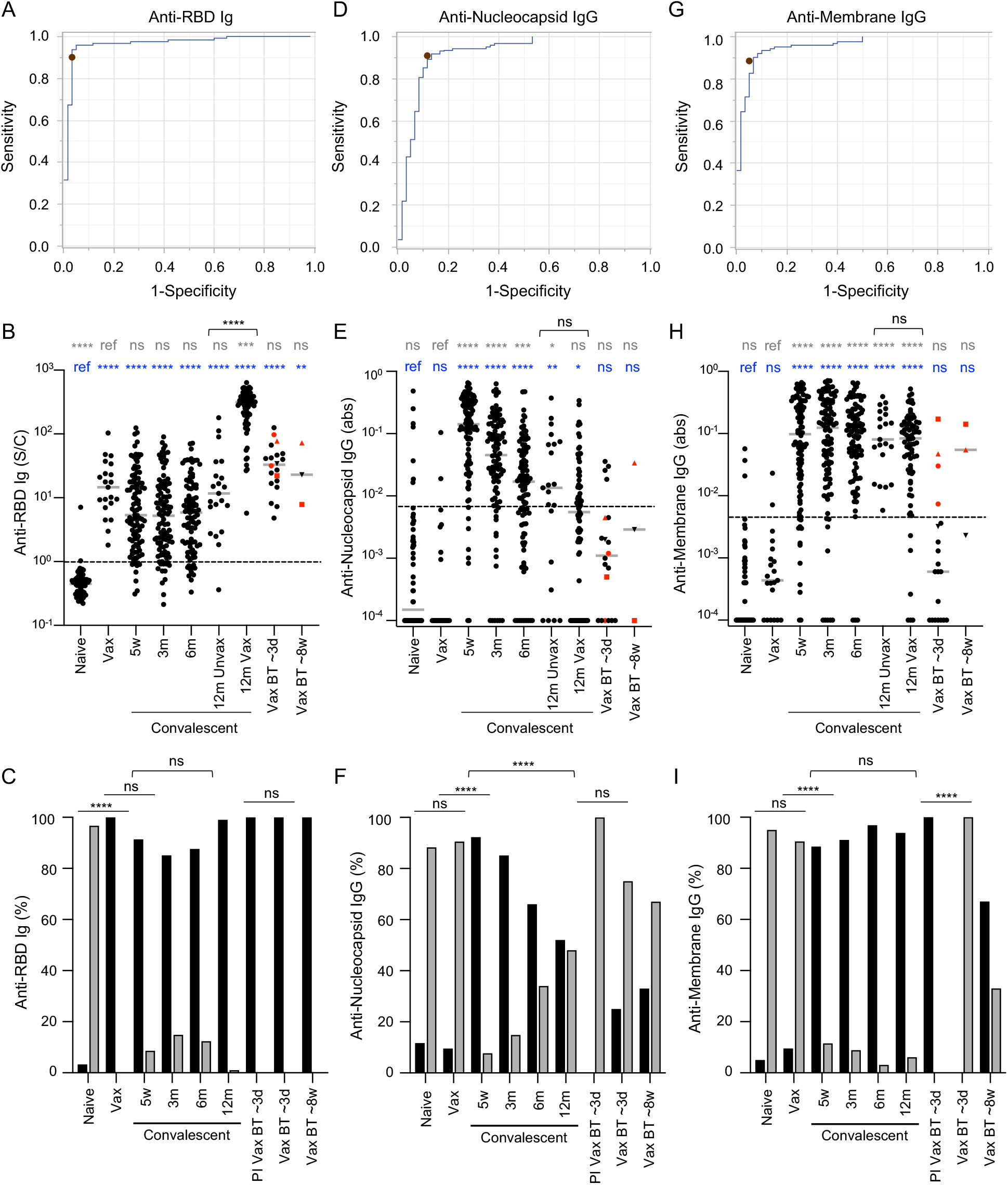
Anti-RBD, anti-nucleocapsid, and anti-membrane antibodies after COVID-19 vaccination and infection. Anti-RBD Ig was detected by immunoassay (A-C) and anti-nucleocapsid (D-F) and anti-membrane (G-I) IgG was quantified by ELISA. A, D, G. ROC curves for antibody levels from naive (n=60) and 5 week COVID-19 convalescent (n=121) sera. Dots indicate cutoffs for seropositivity. Sera from the following subjects were tested in all assays: naive (n=60), vaccinated with no known COVID-19 infection (Vax, n=21), COVID-19 convalescent 5 weeks (5w, n=104), 3 months (3m, n=101), 6 months (6m, n=97), and 12 months (12m, n=98) post-symptom resolution either vaccinated (12m Vax, n=77) or not (12m Unvax, n=21), vaccinated with breakthrough COVID-19 ∼3 days (Vax BT ∼3d, n=20) and ∼8 weeks (Vax BT ∼8w, n=3) after symptom onset including 4 subjects with previous COVID-19 infection (PI). B, E, H. Antibody levels (sample/calibrator, S/C; absorbance, abs) for all groups were graphed and compared to naive (blue) or Vax (gray) by Kruskal-Wallis and Dunn’s multiple comparisons tests and 12m Unvax was compared to 12m Vax by Mann-Whitney test (brackets). Gray bars indicate medians and dashed lines indicate cutoffs. Vax BT with both ∼3d and ∼8w timepoints are represented with triangles or squares and Vax BT subjects with PI in red symbols. C, F, I. Percent positive (black) and negative (gray) for each antibody test were graphed and compared between selected groups by Fisher’s exact (line) or chi-square (bracket encompassing compared groups) tests. For all panels: *p<0.05, **p<0.01, ***p<0.001, ****p<0.0001, or not significant (ns).

Next, we evaluated IgG against the SARS-CoV-2 nucleocapsid peptide ^11^. This ELISA was not as strong as the anti-RBD assay, given the AUC of 0.919 (Wald 95% CI 0.879, 0.959), sensitivity of 91%, and specificity of 88% (Figure 1D). The lower accuracy of this test is consistent with previous reports with different anti-nucleocapsid tests and may be due to inconsistent immune responses to SARS-CoV-2 nucleocapsid in COVID-19 convalescent subjects and cross-reactivity with the nucleocapsid of common cold coronaviruses ^3,6,7,14,15^. Nonetheless, as expected, there was no difference in anti-nucleocapsid IgG levels between naïve versus vaccinated subjects or between 12 month COVID-19 convalescent vaccinated versus unvaccinated subjects (Figure 1E). Also as expected, compared to either vaccinated or naïve subjects, anti-nucleocapsid IgG levels were higher in subjects with past COVID-19 (Figure 1E). However, in a paired analysis of convalescent subjects, anti-nucleocapsid IgG fell rapidly over time and was significantly lower at 3 months (median 0.0446, p<0.0001), 6 months (median 0.0143, p<0.0001) and 12 months (median 0.0071, p<0.0001) compared to 5 weeks (median 0.1417) post-symptom resolution. Consistent with the fall in median levels, 34% of subjects were seronegative for anti-nucleocapsid IgG by 6 months and 48% by 12 months, a significant increase in seronegativity compared to 8% at 5 weeks (Figure 1F). Interestingly, none of the four breakthrough cases who also had COVID-19 before vaccination were seropositive for anti-nucleocapsid IgG at the time of breakthrough infection (Figure 1F). Further, only one of three subjects was seropositive for anti-nucleocapsid IgG eight weeks after breakthrough infection (Figure 1F). Together, these data highlight the rapid decline of anti-nucleocapsid antibodies and the lack of specificity for this antigen.

Lastly, we evaluated IgG against the SARS-CoV-2 membrane peptide ^11^. Our ELISA had an AUC of 0.956 (Wald 95% CI 0.926, 0.986) with a sensitivity of 88% and specificity of 95% (Figure 1G). As expected, anti-membrane IgG levels and percent seropositivity did not differ between naïve and vaccinated subjects or between 12 month convalescent vaccinated and unvaccinated subjects, but were significantly higher in convalescent subjects (Figure 1G and 1H). In a paired analysis over time, anti-membrane IgG levels remained stable at 6 months, with a statistically significant (p=0.0020) decline from 6 months (median 0.1077) to 12 months (median 0.0813). However, at 12 months, 94% of convalescent samples were seropositive for anti-membrane IgG, as compared to 88% seropositive at 5 weeks (Figure 1I). Interestingly, all four vaccine breakthrough infection subjects who had previous COVID-19 were seropositive for anti-membrane IgG during acute infection, while no breakthrough subjects without prior COVID-19 had detectable anti-membrane antibodies in the acute infection period (Figure 1I). Together, these data demonstrate that anti-membrane IgG is a sensitive and specific marker of past COVID-19 infection.

In this study, in addition to confirming that anti-RBD antibodies are long-lasting and anti-nucleocapsid IgG declines rapidly ^3-7^, we show for the first time that anti-membrane IgG is present in the vast majority of COVID-19 convalescent patients and persists for at least a year. Our findings are consistent with findings for IgG against a recombinant membrane antigen (polypeptide of aa 1-19 and 101-222) in the early convalescent period ^12^. In contrast, a separate study found that only ∼20% of non-hospitalized COVID-19 convalescent subjects had IgG against a similar membrane peptide (aa 1-20) to ours (aa 8-23) in the early convalescent period ^13^. Our study included subjects with a wide range of COVID-19 disease severity, but when we evaluated non-hospitalized subjects alone, we found 88% positivity for anti-membrane IgG at 5 weeks and 96% at 12 months post-symptom resolution, suggesting long-lasting antibody detection with our peptide even among subjects without severe disease. Further, although sample size was small, the anti-membrane antibody test detected past COVID-19 infection in individuals with breakthrough infections, whereas the anti-nucleocapsid antibody test did not. Thus, anti-membrane IgG could serve as a sensitive and specific marker of past COVID-19 infection.

This study, which uses multiple different patient subsets including naïve, longitudinally collected COVID-19 convalescent with a wide range of disease severity, vaccinated, and vaccinated with breakthrough COVID-19, suggests that a combination of anti-RBD and anti-membrane antibody testing could be used to detect past infection and vaccination for population-wide surveillance or individual patient care. An analogous important testing strategy is used for hepatitis B, in which antibodies against the surface antigen, which have neutralizing capacity ^16^ like anti-RBD antibodies ^17^, indicate either past infection or vaccination and antibodies against the core protein are only present post-infection. Such testing is critical for identifying individuals who were infected by hepatitis B and might need increased surveillance or prophylactic treatment when immunosuppressed ^18^. While SARS-CoV-2 does not appear to cause persistent infection like hepatitis B, the long term consequences of COVID-19 are still emerging and diagnosing past infection may be critical, a particular challenge if viral testing was not performed during acute infection. Population-wide testing for anti-membrane and anti-spike antibodies may also prove crucial for prevalence assessments and for public health policy if meaningful differences in long-term protective immunity due to infection versus vaccination are revealed.

In addition to demonstrating the longevity and utility of anti-membrane antibodies, our work provides some insight into breakthrough infections, although sample sizes are very small. In contrast to the reduced neutralizing titers reported in vaccinated individuals with breakthrough infections ^19^, we detected similar anti-RBD Ig levels in vaccinated subjects with or without breakthrough infections. Possible causes for this discrepancy are the different antibody assays used, sample size, and our collection of breakthrough infection samples ∼3 days after symptom onset and thus likely ∼7 days post-infection, possibly time for a memory or new primary immune response to begin to raise antibody levels. However, the 4 breakthrough infection subjects who also had COVID-19 prior to vaccination had anti-RBD and anti-nucleocapsid antibody levels at the low end of expected (based on antibody levels in 12 month convalescent patients who were vaccinated), suggesting a reduced antibody response to the original infection. Interestingly, antibodies generally did not rise against any antigen in the three longitudinally followed subjects several weeks after breakthrough infection. Although none of these subjects had immunocompromising conditions, it is not known if the lack of antibody response was due to an atypical host immune system or the effects of prior vaccination on the subsequent immune response to infection. Of note, the breakthrough infections were caused by similar SARS-CoV-2 lineages as generally infecting the community (Supplementary Figure 2). Thus, while we did not identify anti-RBD levels that could predict breakthrough infection, our limited data suggest that individuals with breakthrough infections may mount a reduced antibody response against SARS-CoV-2.

In sum, this study demonstrates that anti-RBD and anti-membrane antibodies are long-lasting and, in combination, can accurately identify past-COVID-19 infection and vaccination in a variety of populations.

## Methods

### Human subjects

Human studies were performed according to the Declaration of Helsinki and were approved by the University of Wisconsin (UW) Institutional Review Board. Sera and data collected prior to 2019 from COVID-19 naïve adults without inflammatory disease (except one subject with psoriatic arthritis using adalimumab to match two COVID-19 convalescent subjects using adalimumab) were obtained from the University of Wisconsin (UW) Rheumatology Biorepository, described in ^20^. COVID-19 convalescent human sera and data were obtained from the UW COVID-19 Convalescent Biorepository, as described in ^14^. Briefly, in the spring of 2020, adults who tested positive by PCR for SARS-CoV-2 at UW Health were invited to participate in the COVID-19 Convalescent Biorepository until 121 subjects were recruited. Demographic and clinical information were collected by questionnaire and abstraction of the electronic medical record. COVID-19 severity was diverse with mild (n=12), moderate (n=86), severe (n=15) and critical (n=8) clinical phenotypes represented as previously defined ^14^. Subjects provided blood and clinical information 5 weeks (n=121), 3 months (n=115), 6 months (n=98) and 12 months (n=100) after symptom resolution. A single 3 month sample collected >3 weeks from the intended timepoint and subjects who had the 5 week time point collected >3 weeks from the intended timepoint (n=1) or missed more than one blood draw (n=16) were excluded from comparative analyses. Subjects that provided samples at all 4 timepoints (n=88) were included in paired longitudinal antibody analyses. Based on the timing of anti-RBD Ig elevation post-vaccination (Supplementary Figure 1), COVID-19 convalescent subjects at the 12 month timepoint were considered vaccinated if they received their first or only vaccine dose at least 5 days before sample collection and unvaccinated if they received no vaccine (n=17) or their first or only vaccine dose (n=4) <5 days before sample collection. Vaccinated individuals with no known COVID-19 (n=21) were recruited by flyers posted at UW Health locations, provided a blood sample, and completed a questionnaire. Vaccination completion and lack of known COVID-19 was confirmed by questionnaire and review of the electronic medical record. Finally, limited clinical data, sera, and SARS-CoV-2 lineages were provided by UW Health Infection Control for 20 fully vaccinated healthcare workers with breakthrough COVID-19 (positive PCR test and symptoms). Blood collection occurred on average one day after the PCR test (range 0-4 days) and on average three days after symptom onset (range 0-5 days). Four of the breakthrough cases also had PCR positive COVID-19 infection prior to the completion of vaccination (3-6 months prior to breakthrough infection). All 20 healthcare workers with breakthrough infections were invited to participate in the longitudinal study, but only 3 consented to provide an additional blood sample ∼8 weeks (range 37-70 days) after the initial collection. According to the electronic medical record, these three subjects were not immunocompromised, defined as no immunosuppressing medications, systemic inflammatory or autoimmune disease, cancer not in remission, uncontrolled diabetes, or congenital or acquired immunodeficiency.

### Anti-membrane and anti-nucleocapsid IgG ELISA

ELISAs were performed as previously described to detect IgG against SARS-CoV-2 membrane (aa 8-23, ITVEELKKLLEQWNLV-K-biotin) and nucleocapsid (aa 390-405, QTVTLLPAADLDDFSK-K-biotin) peptides ^11^ with the following modifications: blocking for >2.5 hours instead of 1 and human sera were diluted 1:50 or 1:500 for nucleocapsid or membrane peptides, respectively, instead of 1:200. Absorbance values of 0 were plotted as 0.0001 to allow a log scale for graphs. Cutoff absorbance values for optimal sensitivity and specificity were 0.0067 for anti-nucleocapsid and 0.0043 for anti-membrane IgG.

### SARS-CoV-2 Immunoassay

Anti-RBD Ig was detected by Lumit^−^ SARS-CoV-2 Immunoassay (Promega, Madison, USA) according to kit instructions using a TEMPEST^®^ Liquid Handler (Formulatrix, Bedford, MA) with luminescence detected using a PHERAstar FS plate reader (BMG Labtech, Ortenberg, Germany). To use the kit’s sample/calibrator cutoff of 1 for seropositivity, sera were diluted 1:10. Although this dilution resulted in samples with the highest anti-RBD Ig values falling outside of the linear range, results were similar at a 1:200 dilution, which captured the linear range for higher samples (Supplementary Figure 3).

### Statistical analysis

Graphing and statistical analyses were performed using Prism (GraphPad, San Diego, CA) and JMP (SAS Institute, Cary, NC) software. Antibody levels were compared between naïve or vaccinated subjects versus all other groups by Kruskal-Wallis One-Way ANOVA with Dunn’s multiple comparisons test. Antibody levels in unvaccinated versus vaccinated 12 month convalescent samples were compared by Mann-Whitney U test. Antibody levels in paired samples from the same convalescent subjects were compared by Friedman test with Dunn’s multiple comparisons test. Wilcoxon signed rank test was used to compare antibody levels between the same subjects with breakthrough infections that provided samples ∼3 days post-symptom onset and at follow-up ∼8 weeks later. Positive antibody detection was compared between two groups with Fisher’s exact test and among multiple groups with a chi-square test. P values <0.05 were considered significant. Data for all subjects is included as a supplemental file.

## Data Availability

All data produced in the present work are contained in the supplemental files.

## Acknowledgements

The authors would like to thank Gene Ananiev and Song Guo at the University of Wisconsin (UW)-Madison’s Small Molecules Screening Facility and Laura Perez-Brenner, Samantha Lewis and Melanie Dart from Promega (Madison, WI) for antibody assay technical advice, UW Health Infection Control and Employee Health Services, the UW School of Medicine and Public Health (UWSMPH) Clinical Research Office, and the human subjects who participated in this research. This work was supported by a Wisconsin Partnership Program COVID-19 Response grant (4647) and a UWSMPH Department of Medicine COVID-19 Pilot Award to MAS, by the Centers for Disease Control (75D30120C09870 and 75D30121C11060) to DHO and TCF, and by the National Institutes of Health (NIH) National Institute of Allergy and Infectious Diseases (R24OD017850) to DHO. Additional support includes NIH National Institute on Aging (T32 AG000213) and Medical Scientist Training Program (T32 GM140935) to MFA, National Heart, Lung and Blood Institute (T32 HL007899) to AMM, National Center for Advancing Translational Sciences (1KL2TR002374) to SSM, and National Institute of Allergy and Infectious Diseases (1DP2AI144244-01) to NS. DHO and TCF are also members of the Upper Midwest Regional Accelerator for Genomic Surveillance funded by the Rockefeller Foundation.

## Author contributions

MAS conceived of, MFA and MAS designed, and DHO, TCF, and MAS oversaw the study. MFA, RRA, SG, SJB, AMM, NS, and MAS participated in subject recruitment, sample collection, and/or sample processing. MFA, KMB, and GKM acquired and MFA, SSM, and MAS analyzed data. MFA and MAS prepared figures and wrote the manuscript. All authors edited the manuscript and approved the final version.

## References

1 Wijeratne, T. & Crewther, S. Post-COVID 19 Neurological Syndrome (PCNS); a novel syndrome with challenges for the global neurology community. J Neurol Sci 419, 117179, doi:10.1016/j.jns.2020.117179 (2020).

2 EUA Authorized Serology Test Performance. https://www.fda.gov/medical-devices/coronavirus-disease-2019-covid-19-emergency-use-authorizations-medical-devices/eua-authorized-serology-test-performance.

3 Ripperger, T. J. et al. Orthogonal SARS-CoV-2 Serological Assays Enable Surveillance of Low-Prevalence Communities and Reveal Durable Humoral Immunity. Immunity 53, 925-933.e924, doi:10.1016/j.immuni.2020.10.004 (2020).

4 Liu, A., Li, Y., Peng, J., Huang, Y. & Xu, D. Antibody responses against SARS-CoV-2 in COVID-19 patients. J Med Virol, doi:10.1002/jmv.26241 (2020).

5 Bolotin, S. et al. SARS-CoV-2 Seroprevalence Survey Estimates Are Affected by Anti-Nucleocapsid Antibody Decline. J Infect Dis 223, 1334–1338, doi:10.1093/infdis/jiaa796 (2021).

6 Gallais, F. et al. Evolution of antibody responses up to 13 months after SARS-CoV-2 infection and risk of reinfection. EBioMedicine 71, 103561, doi:10.1016/j.ebiom.2021.103561 (2021).

7 Wang, Z. et al. Naturally enhanced neutralizing breadth against SARS-CoV-2 one year after infection. Nature 595, 426–431, doi:10.1038/s41586-021-03696-9 (2021).

8 Wang, Z. et al. mRNA vaccine-elicited antibodies to SARS-CoV-2 and circulating variants. Nature 592, 616–622, doi:10.1038/s41586-021-03324-6 (2021).

9 Doria-Rose, N. et al. Antibody Persistence through 6 Months after the Second Dose of mRNA-1273 Vaccine for Covid-19. N Engl J Med 384, 2259–2261, doi:10.1056/NEJMc2103916 (2021).

10 Sadoff, J. et al. Safety and Efficacy of Single-Dose Ad26.COV2.S Vaccine against Covid-19. N Engl J Med 384, 2187–2201, doi:10.1056/NEJMoa2101544 (2021).

11 Heffron, A. S. et al. The landscape of antibody binding in SARS-CoV-2 infection. PLoS Biol 19, e3001265, doi:10.1371/journal.pbio.3001265 (2021).

12 Lopandić, Z. et al. IgM and IgG Immunoreactivity of SARS-CoV-2 Recombinant M Protein. Int J Mol Sci 22, doi:10.3390/ijms22094951 (2021).

13 Jörrißen, P. et al. Antibody Response to SARS-CoV-2 Membrane Protein in Patients of the Acute and Convalescent Phase of COVID-19. Front Immunol 12, 679841, doi:10.3389/fimmu.2021.679841 (2021).

14 Amjadi, M. F. et al. Specific COVID-19 Symptoms Correlate with High Antibody Levels against SARS-CoV-2. Immunohorizons 5, 466–476, doi:10.4049/immunohorizons.2100022 (2021).

15 Noda, K. et al. A novel highly quantitative and reproducible assay for the detection of anti-SARS-CoV-2 IgG and IgM antibodies. Sci Rep 11, 5198, doi:10.1038/s41598-021-84387-3 (2021).

16 Huang, C. F., Lin, S. S., Ho, Y. C., Chen, F. L. & Yang, C. C. The immune response induced by hepatitis B virus principal antigens. Cell Mol Immunol 3, 97–106 (2006).

17 Yu, F. et al. Receptor-binding domain-specific human neutralizing monoclonal antibodies against SARS-CoV and SARS-CoV-2. Signal Transduct Target Ther 5, 212, doi:10.1038/s41392-020-00318-0 (2020).

18 Buch, M. H. et al. Updated consensus statement on the use of rituximab in patients with rheumatoid arthritis. Ann Rheum Dis 70, 909–920, doi:10.1136/ard.2010.144998 (2011).

19 Bergwerk, M. et al. Covid-19 Breakthrough Infections in Vaccinated Health Care Workers. N Engl J Med, doi:10.1056/NEJMoa2109072 (2021).

20 Holmes, C. L. et al. Reduced IgG titers against pertussis in rheumatoid arthritis: Evidence for a citrulline-biased immune response and medication effects. PLoS One 14, e0217221, doi:10.1371/journal.pone.0217221 (2019).

