## Supplementary for "Anti-membrane and anti-spike antibodies are long-lasting and together discriminate between past COVID-19 infection and vaccination"

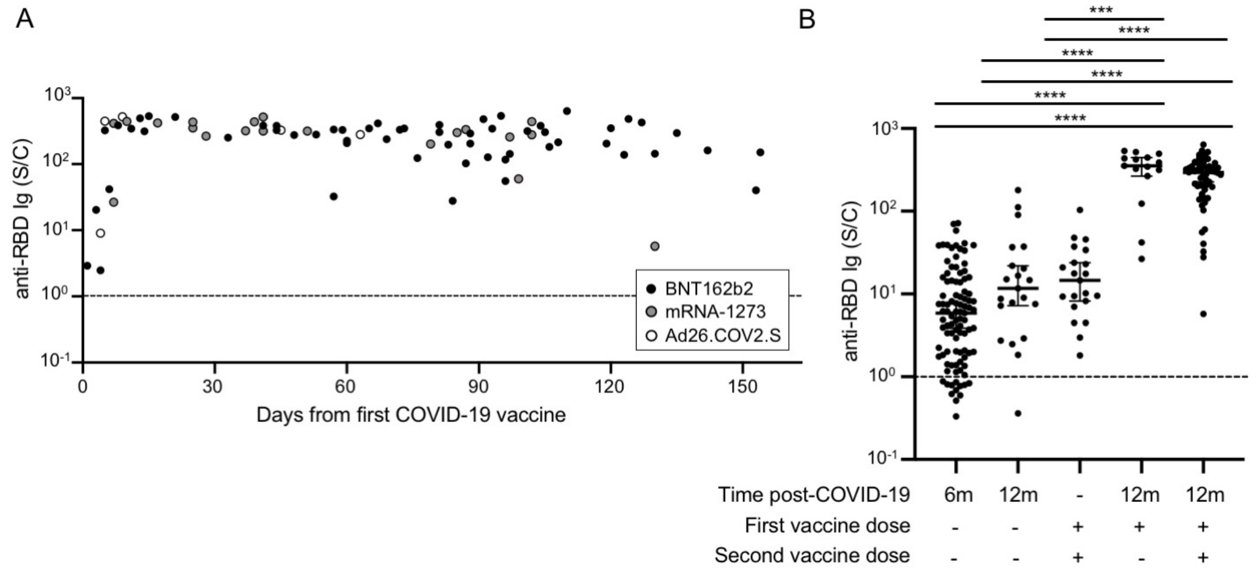


**Supplementary Figure 1. Rise in anti-RBD Ig in response to vaccination in COVID-19 convalescent subjects.** A. Serum anti-RBD Ig levels (reported as sample/calibrator, S/C) for COVID-19 convalescent individuals 12 months after symptom resolution were plotted according to the time of receipt of the first BNT162b2 vaccine dose (n=58), the first mRNA-1273 vaccine dose (n=20), or the only Ad26.COV2.S vaccine dose (n=5). B. Serum anti-RBD Ig levels for unvaccinated COVID-19 convalescent individuals 6 months (n=97) or 12 months (n=21) post symptom resolution, fully vaccinated individuals with no known COVID-19 (n=21), and 12 month convalescent subjects who received either 1 (n=15) or 2 (n=58) vaccine doses of BNT162b2 or mRNA-1273 were compared by Kruskal-Wallis One-way ANOVA with Dunn's multiple comparisons test (***p<0.001, ****p<0.0001). Median and 95% CI are shown. For all panels, black dashed lines indicate the antibody detection cutoff.

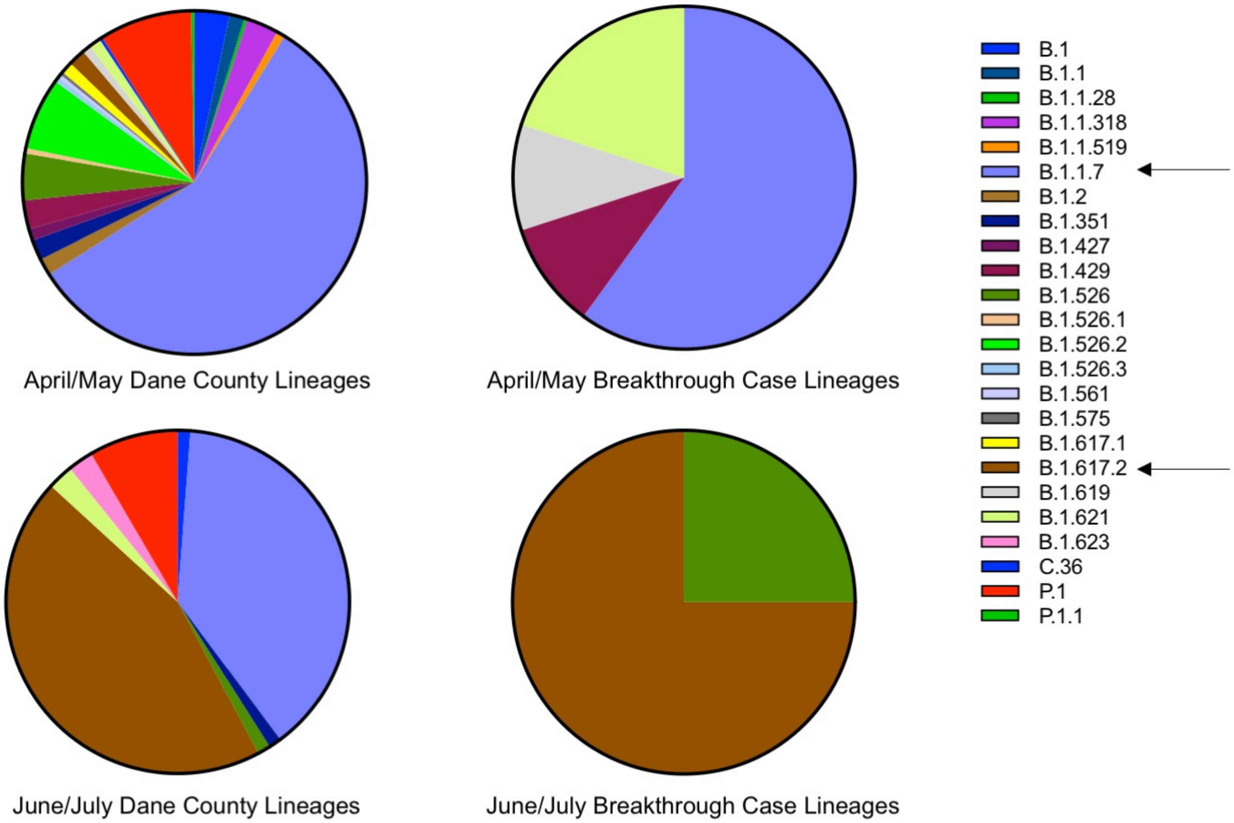


**Supplementary Figure 2. Similar SARS-CoV-2 lineages sequenced in breakthrough and general infections in Dane County.** Samples from PCR positive COVID-19 cases in 2021 in Dane County (n=367 April-May, n=83 June-July) and vaccinated subjects with breakthrough COVID-19 (n=10 April-May, n=4 June-July), who were also located in Dane County, were sequenced to determine viral lineages. Pie charts show percentage of each lineage sequenced out of the total sequenced for each group. Arrows indicate the most common lineages. Six of twenty breakthrough samples could not be sequenced. Of note, B.1.1.7 is commonly called the alpha variant and B.1.617.2 is commonly called the delta variant.

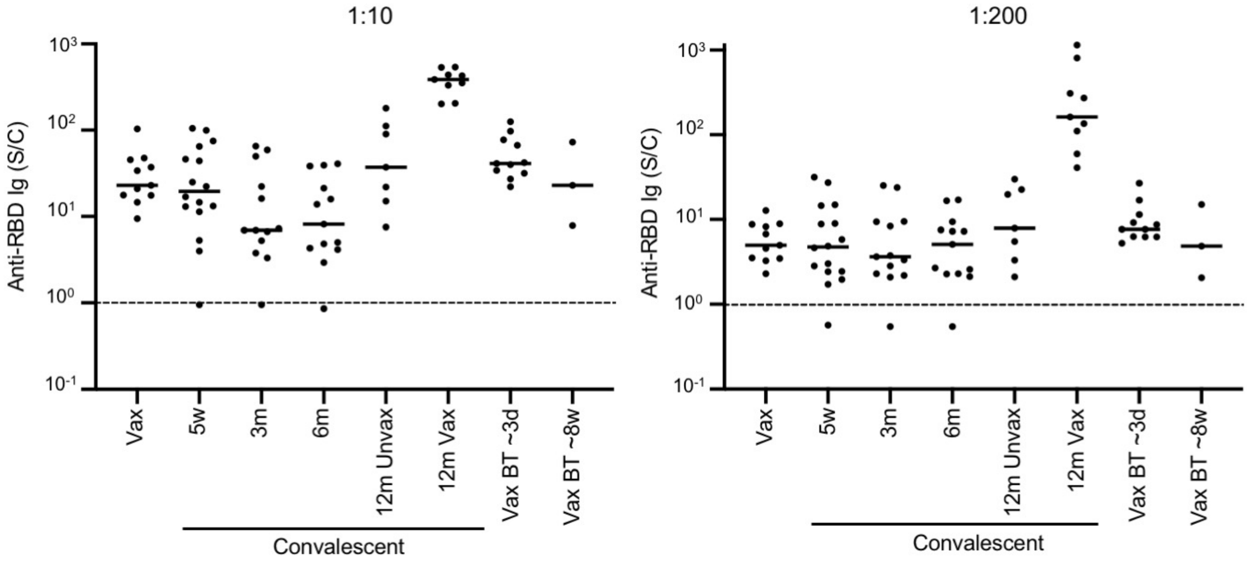


**Supplementary Figure 3. Similar results for anti-RBD Ig detection at different serum dilutions.** Anti-RBD Ig levels (sample/calibrator, S/C) detected by immunoassay in sera from COVID-19 vaccinated subjects with no known COVID-19 infection (Vax n=11), COVID-19 convalescent patients collected 5 weeks (5w, n=16), 3 months (3m, n=13), 6 months (6m, n=13), and 12 months post symptom resolution (n=7 unvaccinated, 12m Unvax; n=9 vaccinated, 12m Vax), vaccinated subjects with breakthrough COVID-19 (n=11) collected ~3 days (Vax BT ~3d) and ~8 weeks (Vax BT ~8w, n=3) after symptom onset showed similar trends when evaluated at 1:10 and 1:200 sera dilutions. Black solid lines indicate medians and black dashed lines represent antibody detection cutoffs.
